# Clinical characteristics and outcomes of COVID-19 breakthrough infections among vaccinated patients with systemic autoimmune rheumatic diseases

**DOI:** 10.1101/2021.08.04.21261618

**Authors:** Claire Cook, Naomi J. Patel, Kristin M. D’Silva, Tiffany Y-T. Hsu, Michael DiIorio, Lauren Prisco, Lily Martin, Kathleen M.M. Vanni, Alessandra Zaccardelli, Derrick J. Todd, Jeffrey A. Sparks, Zachary S. Wallace

**Author notes:** Contributed equally (Jeffrey A. Sparks, Zachary S. Wallace). **<u>Corresponding author:</u>** Zachary S. Wallace, MD, MSc, Clinical Epidemiology Program, Division of Rheumatology, Allergy, and Immunology, Massachusetts General Hospital, 100 Cambridge Street, 16th Floor, Boston, MA 02114, @zach_wallace_md. **<u>Competing interests:</u>** JAS reports research support from Bristol-Myers Squibb and consultancy fees from Bristol-Myers Squibb, Gilead, and Pfizer. ZSW reports research support from Bristol-Myers Squibb and Principia/Sanofi and consulting fees from Viela Bio and MedPace. All other authors report no competing interests.

## Abstract

**Objective:** To describe the characteristics of COVID-19 vaccine breakthrough infections among systemic autoimmune rheumatic disease (SARD) patients.

**Methods:** We identified SARDs patients in a large healthcare system with COVID-19 vaccination ≥14 days prior to a positive SARS-CoV-2 molecular test. Details of the patient’s SARD, vaccination status, and COVID-19 infection were extracted.

**Results:** Of 340 confirmed COVID-19 infections among SARDs patients between December 11^th^, 2020 (date of first COVID-19 vaccine approval in the US) and July 30^th^, 2021, we identified 16 breakthrough infections. Seven (44%) received the Pfizer-BioNtech vaccine, five (31%) received the Moderna vaccine, and four (25%) received the Janssen/Johnson & Johnson vaccine. The most common SARDs included rheumatoid arthritis (6, 38%), inflammatory myopathy (3, 19%), and systemic lupus erythematosus (3, 19%). Rituximab (5, 31%), glucocorticoids (4, 25%), and mycophenolate mofetil (4, 25%) were the most frequent treatments. Among the breakthrough infections, 15 (93%) were symptomatic, six (38%) were hospitalized, one (6%) required mechanical ventilation, and two (13%) died.

**Conclusions:** Symptomatic, including severe, breakthrough infections were observed in SARDs patients; many were on treatments associated with attenuated antibody responses to vaccination. Further studies are needed to determine the rate of breakthrough infection associated with SARD treatments and other features.

**Key messages:** *What is already known about this subject?:* Breakthrough infections following COVID-19 vaccination are expected but some patients with systemic autoimmune rheumatic diseases (SARDs) may be at higher risk because of blunted antibody responses to vaccination associated with rheumatic disease treatments and other factors that remain poorly understood.

*What does this study add?:* We identify and describe 16 COVID-19 vaccine breakthrough infections within the Mass General Brigham system between December 11^th^, 2020 and June 26^th^, 2021. The vast majority of cases were symptomatic and two were fatal.

*How might this impact on clinical practice or future developments?:* This study complements observations regarding the attenuated antibody response to COVID-19 vaccination in patients with SARDs by identifying serious clinical outcomes from breakthrough infections in patients receiving DMARDs that have been reported to have blunted vaccine responses. Our study identifies characteristics of COVID-19 breakthrough infections that may guide the prioritization of booster vaccines and other risk-mitigating strategies in patients with SARDs.

## INTRODUCTION

SARS-CoV-2 vaccines, including the three with emergency use authorization in the United States (BNT162b2 [Pfizer/BioNTech], mRNA-1273 [Moderna], and AD26.COV2.S [Janssen/Johnson & Johnson]), reduce the risk of COVID-19 (1-3). Patients with systemic autoimmune rheumatic diseases (SARDs) were excluded from the initial COVID-19 vaccine trials. Thus, vaccine efficacy in patients with SARDs, many of whom use disease modifying anti-rheumatic drugs (DMARDs), is unknown. Early reports indicate that certain DMARD use, particularly glucocorticoids, methotrexate, mycophenolate mofetil, and rituximab, may blunt the immunologic response to COVID-19 vaccination (4-7). However, little is known about the clinical efficacy of these vaccines at preventing COVID-19 infection in patients with SARDs. We systematically identified breakthrough COVID-19 infections in a large healthcare system to characterize these cases and identify SARDs patients in whom clinical vaccine efficacy may be reduced.

## METHODS

### Data source and study population

Mass General Brigham (MGB) is a large multicenter health care system in the Boston, Massachusetts, USA area that includes tertiary care hospitals, community hospitals, and outpatient clinics. We utilized the MGB centralized data warehouse to identify patients who were ≥18 years of age and had positive polymerase chain reaction (PCR) testing for SARS-CoV-2 between January 30^th^, 2020 and July 30^th^, 2021. Patients with SARDs with a positive SARS-CoV-2 test were identified using diagnostic billing codes, as previously described (8-12). Additional cases with positive PCR or SARS-CoV-2 antigen test results were identified via physician referral from the MGB rheumatology clinics. Electronic health records of identified cases were manually reviewed to confirm the SARD diagnosis and gather details of COVID-19 infection, vaccination, and outcome. Patients with any history of COVID-19 vaccination prior to their COVID-19 molecular test were identified from this cohort. Breakthrough infections in fully vaccinated patients were considered to be those who had SARS-CoV-2 detected by either PCR or antigen testing at least 14 days after their final vaccine dose, as defined by the US Centers for Disease Control and Prevention (13). Partially vaccinated patients were considered to be those with a positive test between 7 and 14 days after their final dose. Final vaccination dose was considered either the first dose in a one-dose series (Janssen/J&J) or second dose in a two-dose vaccination series (Pfizer-BioNTech and Moderna). Descriptive statistics (mean, standard deviation, frequencies, proportions) were used.

### Patient and Public Involvement

Patients or the public were not involved in the design, conduct, reporting, or dissemination of this study.

## RESULTS

### Patient Characteristics

Of 786 SARDs patients with COVID-19 infection identified within the MGB healthcare system, 340 occurred after the initial emergency use authorization for COVID-19 vaccination in the US (December 11^th^, 2020). Of these, 40 (12%) received a COVID-19 vaccination prior to their earliest positive test, 16 (4.7%) of whom were classified as breakthrough infections (**Figure 1**).

Among the 16 breakthrough infections, 12 (75%) were female, 11 (69%) were white, and the median age was 50 (IQR: 38, 64) years (**Table 1**). Twelve patients (75%) had at least one comorbidity. The most common SARDs included rheumatoid arthritis (6, 38%), inflammatory myositis (3, 19%), and systemic lupus erythematosus (3, 19%). Rituximab (5, 31%), glucocorticoids (5, 31%), mycophenolate mofetil or mycophenolic acid (4, 25%), and methotrexate (3, 19%) were the most frequent immunosuppressive medications recorded prior to first vaccine dose. In one case (6%), the patient was on no DMARD or glucocorticoid at the time of their COVID-19 vaccine.

**Table 1.** Patient characteristics, vaccination details, medication use, and infection details of COVID-19 breakthrough infections in fully vaccinated SARDs patients (N=16)

| **Patient Characteristics** n, % |  |
| --- | --- |
| Female | 12, 75 |
| Age (median, IQR) | 49.5 [38.0, 64.5] |
| Race |  |
| White | 11, 68 |
| Black | 3, 20 |
| Hispanic | 2, 13 |
| Rheumatic disease* |  |
| Rheumatoid arthritis | 6, 38 |
| Rheumatoid arthritis-associated interstitial lung disease | 1, 6 |
| Dermatomyositis | 3, 19 |
| Myositis-associated interstitial lung disease | 1, 6 |
| Systemic lupus erythematosus | 3, 19 |
| Ankylosing spondylitis | 2, 13 |
| IgG4-related disease | 1, 6 |
| Mixed connective tissue disease | 1, 6 |
| Hypocomplementemic urticarial vasculitis | 1, 6 |
| Psoriatic arthritis | 1, 6 |
| Comorbidities* |  |
| Hypertension | 6, 38 |
| Morbid obesity (BMI $\geq 40.0$ kg/m<sup>2</sup>) | 3, 19 |
| Interstitial lung disease | 2, 13 |
| End stage renal disease | 2, 13 |
| Chronic Obstructive Pulmonary Disease | 1, 6 |
| Asthma | 1, 6 |
| Diabetes | 1, 6 |
| Obesity (BMI $\geq 30.0$ kg/m<sup>2</sup>) | 1, 6 |
| Coronary artery disease | 1, 6 |
| Cancer | 1, 6 |
| Organ transplant | 1, 6 |
| Immunodeficiency | 1, 6 |
| Chronic neurological or neuromuscular disease | 1, 6 |
| Inflammatory bowel disease | 1, 6 |
| None | 4, 25 |
| Smoking status |  |
| Current | 1, 6 |
| Former | 7, 44 |
| Never | 6, 38 |
| Unknown | 2, 13 |
| **Vaccination Details** n, % |  |
| Vaccine Type |  |
| Pfizer-BioNtech | 7, 44 |
| Moderna | 5, 31 |
| Janssen/Johnson & Johnson | 4, 25 |
| Disease Activity at Vaccination |  |
| First vaccination |  |
| Active | 5, 31 |
| Inactive | 11, 69 |
| Second vaccination |  |
| Active | 6, 38 |
| Inactive | 6, 38 |
| Not applicable | 4, 25 |
| **Rheumatic Disease Treatment Use Prior to First Vaccine Dose<sup>†</sup>** n, % |  |
| Rituximab | 5, 31 |
| Glucocorticoids | 5, 31 |
| Mycophenolate mofetil or mycophenolic acid | 4, 25 |
| Methotrexate | 3, 19 |
| Tacrolimus | 2, 13 |
| Adalimumab | 1, 6 |
| Azathioprine | 1, 6 |
| Belimumab | 1, 6 |
| Hydroxychloroquine | 1, 6 |
| Intravenous Immunoglobulin | 1, 6 |
| Sulfasalazine | 1, 6 |
| Tocilizumab | 1, 6 |
| Ustekinumab | 1, 6 |
| None | 1, 6 |
| **COVID-19 Infection Details** n, % |  |
| Time (days) from second/final vaccine dose to infection (median, IQR) | 54.0 [29.8, 79.0] |
| Infection acquisition |  |
| Close contact with confirmed or probable case of COVID-19 | 4, 25 |
| Presence in a healthcare facility with COVID-19 cases | 3, 19 |
| Community acquired | 3, 19 |
| Unknown | 8, 50 |
| Treatment |  |
| No treatment/supportive care only | 7, 44 |
| Remdesivir | 6, 38 |
| Glucocorticoids | 3, 19 |
| Neutralizing monoclonal antibody | 4, 25 |
| Azithromycin | 2, 13 |
| Convalescent plasma | 1, 6 |
| Enrolled in clinical trial<sup>‡</sup> | 1, 6 |
| Any Symptoms |  |
| Yes | 15, 93 |
| No<sup>§</sup> | 1, 6 |
| Symptoms |  |
| Fever | 9, 56 |
| Cough | 7, 44 |
| Malaise | 6, 38 |
| Myalgia | 5, 31 |
| Rhinorrhea | 5, 31 |
| Headache | 4, 25 |
| Shortness of breath | 4, 25 |
| Sore throat | 4, 25 |
| Diarrhea/vomiting/nausea | 3, 19 |
| Anosmia | 3, 19 |
| Dysgeusia | 3, 19 |
| Chest pain | 2, 13 |
| Arthralgia | 1, 6 |
| Other | 1, 6 |
| **Outcomes n, %** |  |
| Outpatient management alone | 10, 63 |
| Hospitalization | 6, 38 |
| Ventilation | 1, 6 |
| Death | 2, 13 |
| Unresolved symptoms | 2, 13 |

\* Patients may have >1 SARD or comorbidity
† One patient initiated methotrexate in between the first and second dose and one patient initiated rituximab between the second dose and infection
‡ NCT04501978; Phase 3 randomized, blinded, trial assessing treatments for hospitalized patients with COVID-19. Intervention arms: investigational drug + standard of care (Remdesivir) or placebo + standard of care (Remdesivir)
§ Diagnosed via pre-procedure PCR test, no reported symptoms in electronic health record

### COVID-19 Vaccination

Seven (44%) patients received the BNT162b2 (Pfizer-BioNtech) vaccine, five (31%) received the mRNA-1273 (Moderna) vaccine, and four (25%) received the AD26.COV2.S (Janssen/Johnson & Johnson) vaccine. At the time of vaccination, five (31%) patients had active disease and eleven (69%) had inactive disease. The median time from final vaccine dose to infection was 54 (IQR 30, 79) days (**Table 1**).

### COVID-19 Infection Characteristics and Outcomes

Among the 16 patients with breakthrough infections, 15 (93%) were symptomatic; the most frequent symptoms included fever (9, 56%), cough (7, 44%), and malaise (6, 38%) (**Table 1**). One (6%) patient was asymptomatic; testing was performed for pre-procedure screening. Six (38%) patients were hospitalized during which four (25%) required supplemental oxygen and one (6%) required mechanical ventilation (**Table 2**). The hospitalized patients had myositis (2, 13%), systemic lupus erythematosus (2, 13%), and rheumatoid arthritis (2, 13%). DMARDs used prior to infection among hospitalized patients included rituximab (4, 25%) and mycophenolate mofetil or mycophenolic acid (2, 13%).

**Table 2.** Descriptions of COVID-19 breakthrough infections ≥14 days after second/final vaccine dose among patients with systemic rheumatic diseases (n=16)

| Case | Age range | Rheumatic Disease | Vaccine Type | Time from final vaccine to diagnosis (days) | Rheumatic disease treatment at the time of first vaccine dose | Rheumatic disease treatment at the time of second vaccine dose (if different/new from first) | Time from last B cell depletion before vaccination to first vaccine dose (days) | Asymptomatic infection | COVID-19 treatment | Hospitalization | Supplemental oxygen/mechanical ventilation | Deceased |
| --- | --- | --- | --- | --- | --- | --- | --- | --- | --- | --- | --- | --- |
| 1 | 45-64 | Dermatomyositis | Moderna | 16 | Azathioprine, Rituximab |  | 99 | No | Remdesivir, azithromycin, glucocorticoids | Yes | Supplemental oxygen | No |
| 2 | 45-64 | Systemic lupus erythematosus | Pfizer-BioNTech | 24 | Mycophenolate Moferil, Tacrolimus, Prednisone (5mg/d) |  |  | No | Remdesivir, Neutralizing monoclonal antibody | Yes | Supplemental oxygen | No |
| 3 | >85 | Rheumatoid arthritis | Pfizer-BioNTech | 26 | Prednisone (10mg/d) | Methotrexate* |  | No | None | No | No | No |
| 4 | 45-64 | Hypocomplementemic urticarial vasculitis, Mixed connective tissue disease | Pfizer-BioNTech | 29 | Hydroxy-chloroquine, Methotrexate |  |  | No | None | No | No | No |
| 5 | 25-44 | Systemic lupus erythematosus | Moderna | 30 | Mycophenolic Acid, Belimumab |  |  | No | Remdesivir | Yes<sup>†</sup> | No | No |
| 6 | 45-64 | Rheumatoid arthritis | Janssen/Johnson & Johnson | 33 | None |  |  | Yes<sup>‡</sup> | None | No | No | No |
| 7 | 65-84 | Rheumatoid arthritis | Janssen/Johnson & Johnson | 40 | Rituximab |  | 190 | No | Remdesivir, glucocorticoids | Yes | Supplemental oxygen | No |
| 8 | 45-64 | Systemic lupus erythematosus | Pfizer-BioNTech | 50 | Mycophenolic acid, Tacrolimus |  |  | No | Neutralizing monoclonal antibody | No | No | No |
| 9 | 65-84 | Ankylosing spondylitis, | Pfizer-BioNTech | 57 | Rituximab, prednisone (20mg/d) |  | 25 | No | Remdesivir, glucocorticoids, | Yes | Supplemental oxygen and | Yes |
|  |  | Dermatomyositis with interstitial lung disease |  |  |  |  |  |  | Convalescent plasma, Enrolled in clinical trial<sup>§</sup> |  | high-flow oxygen device |  |
| 10 | 25-44 | Dermatomyositis | Moderna | 61 | IVIG, Mycophenolate mofetil, Rituximab |  | 111 | No | None | No | No | No |
| 11 | 25-44 | Rheumatoid arthritis | Pfizer-BioNTech | 70 | Methotrexate |  |  | No | None | No | No | No |
| 12 | 65-84 | Rheumatoid arthritis-associated interstitial lung disease | Moderna | 78 | Rituximab |  | 35 | No | Remdesivir, azithromycin, glucocorticoids | Yes | Mechanical ventilation | Yes |
| 13 | 25-44 | IgG4-related disease | Moderna | 82 | Prednisone (10mg/d)<sup>¶</sup> |  |  | No | None | No | No | No |
| 14 | 18-24 | Rheumatoid arthritis | Pfizer-BioNTech | 96 | Methotrexate, Tocilizumab |  |  | No | Neutralizing monoclonal antibody | No | No | No |
| 15 | 25-44 | Ankylosing spondylitis | Janssen/Johnson & Johnson | 111 | Adalimumab, Sulfasalazine |  |  | No | None | No | No | No |
| 16 | 25-44 | Psoriatic arthritis | Janssen/Johnson & Johnson | 132 | Ustekinumab, prednisone (10mg/day) |  |  | No | Neutralizing monoclonal antibody | No | No | No |

IVIG, Intravenous immunoglobulin
\* Initiated methotrexate between dose one and two
† Hospitalization unrelated to COVID-19 diagnosis
‡ Diagnosed via pre-procedure PCR test, no reported symptoms in electronic health record
§ NCT04501978; Phase 3 randomized, blinded, trial assessing treatments for hospitalized patients with COVID-19. Intervention arms: investigational drug + standard of care (Remdesivir) or placebo + standard of care (Remdesivir)
¶ Initiated rituximab between the second dose and infection

Seven (44%) patients did not receive COVID-19 treatment. Three (19%) received neutralizing monoclonal antibody treatment as outpatients. Six (38%) patients were treated with remdesivir, and three (19%) received glucocorticoids. Two (13%) patients died; both deceased patients had received rituximab and had interstitial lung disease. Symptoms were unresolved (one active infection recent diagnosed, one reporting ongoing symptoms: fatigue/malaise) in two (13%) cases. There was only one breakthrough infection each in patients receiving a tumor necrosis factor inhibitor or tocilizumab, and both were also receiving concomitant conventional DMARDs. Similar trends in patient features, vaccination, and infection characteristics were observed among partially vaccinated patients (**Supplemental Tables 1 and 2**).

## DISCUSSION

A small portion of COVID-19 cases among SARDs patients in a large US healthcare system occurred among fully vaccinated patients. Reassuringly, the majority of these breakthrough infections were non-severe and managed as outpatients. However, some patients required hospitalization that ultimately culminated in death. The most commonly used SARDs treatments at the time of vaccination included those associated with blunted antibody responses to SARS-CoV-2 vaccination, including glucocorticoids, methotrexate, mycophenolate mofetil or mycophenolic acid, and rituximab (4-7). These results have immediate implications for SARDs patients.

In contrast to outcomes of breakthrough infections in the general population reported to the United States Centers for Disease Control and Prevention (n=10,262), we observed a larger proportion of patients who required hospitalization (40% vs. 10%) or died (13% vs. 2%) (13). Outcomes in our cohort were also worse than those described in a study of fully-vaccinated skilled nursing home residents and staff (n=22), of whom 18% were hospitalized and 5% died (14). While our small sample size limits the interpretation of these comparisons, the frequency with which use of DMARDs previously associated with blunted antibody responses were observed in our cohort is notable. Indeed, a study of patients hospitalized with breakthrough infections in Israel found that 40% of these patients were considered immunocompromised (15). However, it is possible that we did not identify SARDs patients with asymptomatic or less severe breakthrough infections.

Collectively, these findings suggest that the blunted SARS-CoV-2 antibody response following COVID-19 vaccination in certain DMARD users may be associated with an increased risk of breakthrough infections that may be severe and even fatal. The protective role of SARS-CoV-2-specific T cell responses following vaccination among DMARD users is an area of active investigation, especially among B cell depleting agent users (4). However, our observation that rituximab was the most commonly used DMARD among patients with breakthrough infections and the DMARD used in both fatal cases, raises concerns regarding the ability to mount a T cell immune response against SARS-CoV-2 after B cell depletion. However, both patients also had severe pre-existing interstitial lung disease which likely also contributed to their poor clinical outcome. Some patients treated with B cell depleting agents may require alternative risk mitigation strategies, including passive immunity or booster vaccines, and may need to continue shielding practices (e.g., face masking, social distancing) as society continues to re-open, especially if they have a poor antibody response to vaccination. Perhaps reassuringly, some DMARDs commonly used to treat SARDs had either few (e.g., tumor necrosis factor inhibitors and interleukin-6 receptor inhibitors) or no (e.g., abatacept and Janus kinase inhibitors) (16) identified breakthrough infections in our case series.

Our study has certain strengths, including the systematic approach to case identification and SARD-specific details available from the EHR. Despite these strengths, our study has certain limitations. First, we could not estimate the rate of breakthrough infections among SARDs patients. Second, this was a retrospective study that identified patients who presented to care for SARS-CoV-2 PCR testing, so the proportion of asymptomatic breakthrough infections observed in our study may be an underestimate. Third, while the observed frequency of certain DMARD use (e.g., mycophenolate mofetil, rituximab) in our cohort suggests that the blunted antibody response associated with these DMARDs may put patients at risk for breakthrough infection, we did not have antibody testing available for all patients and cannot rule out the possibility that SARD manifestations (e.g., interstitial lung disease) commonly treated with these medications contributed to the severity of the presentation. Fourth, we were unable to determine whether DMARDs were held temporarily prior to or after vaccine doses as recommended by the American College of Rheumatology (17) since these changes were not consistently documented in the EHR.

In conclusion, a small minority of COVID-19 cases among SARDs patients in a large healthcare system occurred in fully vaccinated patients. However, a large portion of these cases required hospitalization and occurred among patients using DMARDs that have been previously associated with a blunted antibody response following vaccination. Additional studies are urgently needed to estimate the risk of breakthrough infections among SARDs patients and to evaluate the efficacy of booster vaccines and other strategies for DMARD users with poor immunologic response to COVID-19 vaccination.

## Data Availability

Data is available upon reasonable request. Identifying patient data is unavailable.

