## Supplemental Table for "Clinical characteristics and outcomes of COVID-19 breakthrough infections among vaccinated patients with systemic autoimmune rheumatic diseases"

**Supplemental Table 1.** Patient characteristics, vaccination details, medication use, and infection details of breakthrough COVID-19 infections between ≥7 days and <14 days after final vaccine dose (N=5)

| **Patient Characteristics** n, % |  |
| --- | --- |
| Female | 3, 60 |
| Age (mean, SD) | 57.6, 21.4 |
| Race |  |
| Black | 1, 20 |
| Asian | 1, 20 |
| White | 3, 60 |
| Rheumatic disease^*^ |  |
| Rheumatoid arthritis | 2, 40 |
| Rheumatoid arthritis-associated interstitial lung disease | 1, 20 |
| Polymyalgia rheumatica | 1, 20 |
| Seronegative spondyloarthritis | 1, 20 |
| ANCA-associated vasculitis | 1, 20 |
| Comorbidities^*^ |  |
| Obstructive sleep apnea | 2, 40 |
| Diabetes | 2, 40 |
| Obesity (BMI ≥ 30.0 kg/m^2^) | 2, 40 |
| Hypertension | 2, 40 |
| Interstitial lung disease | 1, 20 |
| Chronic obstructive pulmonary disease | 1, 20 |
| Asthma | 1, 20 |
| Morbid obesity (BMI ≥ 40.0 kg/m^2^) | 1, 20 |
| Cancer | 1, 20 |
| None | 2, 40 |
| Smoking status |  |
| Current | 0, 0 |
| Former | 3, 60 |
| Never | 2, 40 |
| **Vaccination Details** n, % |  |
| Vaccine Type |  |
| Pfizer-BioNtech | 4, 80 |
| Moderna | 1, 20 |
| Janssen/Johnson & Johnson | 0, 0 |
| Disease Activity at Vaccination |  |
| First vaccination |  |
| Active | 3, 60 |
| Inactive | 2, 40 |
| Second vaccination |  |
| Active | 2, 40 |
| Inactive | 2, 40 |
| Unknown | 1, 20 |
| **Rheumatic Disease Treatment Prior to First Vaccine Dose** n, % |  |
| Rituximab | 2, 40 |
| Glucocorticoids | 2, 40 |
| Tofacitinib | 1, 20 |
| Methotrexate | 1, 20 |
| Leflunomide | 1, 20 |
| Azathioprine | 1, 20 |
| None | 1, 20 |
| **Infection Details** n, % |  |
| Time (days) second/final vaccine to infection (median, IQR) | 8 [8, 9] |
| Infection acquisition |  |
| Close contact with confirmed or probably case of COVID-19 infection | 2, 40 |
| Unknown | 3, 60 |
| Treatment at Infection |  |
| Glucocorticoids | 3, 60 |
| No treatment | 2, 40 |
| Remdesivir | 2, 40 |
| Convalescent plasma | 2, 40 |
| Other (Ivermectin) | 1, 20 |
| Any Symptoms |  |
| Yes | 4, 80 |
| No^†^ | 1, 20 |
| Symptoms |  |
| Cough | 4, 100 |
| Shortness of breath | 3, 75 |
| Malaise | 3, 75 |
| Fever | 2, 50 |
| Myalgia | 2, 50 |
| Headache | 1, 25 |
| Sore throat | 1, 25 |
| Chest pain | 1, 25 |
| Diarrhea/vomiting/nausea | 1, 25 |
| Rhinorrhea | 1, 25 |
| Arthralgia | 0, 0 |
| **Outcomes** n, % |  |
| Outpatient management alone | 3, 60 |
| Hospitalization | 2, 40 |
| Ventilation | 2, 40 |
| Death | 1, 20 |
| Unresolved symptoms | 1, 20 |

* Patients may have >1 SARD or comorbidity

† Per rheumatologist note in electronic health record: positive PCR test outside MGB system, asymptomatic

### **Supplemental Table 2.** COVID-19 breakthrough infections between ≥7 days and <14 days after second/final vaccine dose (n=5)

| **Patient** | **Age Range** | **Rheumatic Disease** | **Vaccine Type** | **Time from vaccine completion to COVID-19 diagnosis (days)** | **Rheumatic disease treatment at the time of first vaccine dose** | **Time from last B cell depletion before vaccination to first vaccine dose (days)** | **Asympto-matic**  **infection** | **COVID-19 treatment** | **Hospitali-zation** | **Supplemental oxygen/**  **mechanical ventilation** | **Deceased** |
| --- | --- | --- | --- | --- | --- | --- | --- | --- | --- | --- | --- |
| 1 | 65-84 | Polymyalgia rheumatica | Moderna | 7 | Prednisone (7.5mg/d) |  | No | Glucocorticoids | No | No | No |
| 2 | 65-84 | Rheumatoid arthritis-associated interstitial lung disease | Pfizer-BioNTech | 8 | Azathioprine, Rituximab, prednisone  (5 mg/d) | 144 | No | Remdesivir, Glucocorticoids, Convalescent plasma, ivermectin | Yes | Mechanical ventilation | Yes |
| 3 | 45-64 | Rheumatoid arthritis | Pfizer-BioNTech | 8 | Methotrexate |  | No | None | No | No | No |
| 4 | 45-64 | ANCA-associated vasculitis | Pfizer-BioNTech | 9 | Rituximab | 113 | No | Remdesivir, Glucocorticoids, Convalescent plasma | Yes | Mechanical ventilation | No |
| 5 | 18-24 | Seronegative spondyloarthritis | Pfizer-BioNTech | 12 | Leflunomide, Tofacitinib |  | Yes^*^ | None | No | No | No |

* Per rheumatologist note in electronic health record: positive PCR test outside MGB system, asymptomatic
